# Prenatal maternal infections and early childhood developmental outcomes: Analysis of linked administrative health data for Greater Glasgow & Clyde, Scotland

**DOI:** 10.1101/2023.09.25.23296070

**Authors:** Iain Hardie, Aja Murray, Josiah King, Hildigunnur Anna Hall, Emily Luedecke, Louise Marryat, Lucy Thompson, Helen Minnis, Philip Wilson, Bonnie Auyeung

## Abstract

**Background:** Previous research has linked prenatal maternal infections to later childhood developmental outcomes and socioemotional difficulties. However, existing studies have relied on retrospectively self-reported survey data, or data on hospital-recorded infections only, resulting in gaps in data collection.

**Methods:** This study used a large linked administrative health dataset, bringing together data from birth records, hospital records, prescriptions and routine child health reviews for 55,856 children born in Greater Glasgow & Clyde, Scotland, in 2011-2015, and their mothers. Logistic regression models examined associations between prenatal infections, measured as both hospital-diagnosed prenatal infections and receipt of infection-related prescription(s) during pregnancy, and childhood developmental concern(s) identified by health visitors during 6-8 weeks or 27-30 months health reviews. Secondary analyses examined whether results varied by (a) specific developmental outcome types (gross-motor-skills, hearing-communication, vision-social-awareness, personal-social, emotional-behavioural-attention, and speech-language-communication), and (b) the trimester(s) in which infections occurred.

**Results:** After confounder/covariate adjustment, hospital-diagnosed infections were associated with increased odds of having at least one developmental concern (OR: 1.30; 95% CI: 1.19-1.42). This was consistent across almost all developmental outcome types, and appeared to be specifically linked to infections occurring in pregnancy trimesters 2 (OR: 1.34; 95% CI: 1.07-1.67) and 3 (OR: 1.33; 95% CI: 1.21-1.47), i.e. the trimesters in which fetal brain myelination occurs. Infection-related prescriptions were not associated with a significant increase in odds of having at least one developmental concern after confounders/covariate adjustment (OR: 1.03; 95% CI: 0.98-1.08), but were associated with slightly increased odds of concerns specifically related to personal-social (OR: 1.12; 95% CI: 1.03-1.22) and emotional-behavioural-attention (OR: 1.15; 95% CI: 1.08-1.22) development.

**Conclusions:** Prenatal infections, particularly those which are hospital-diagnosed (and likely more severe) are associated with early childhood developmental outcomes. Prevention of prenatal infections, and monitoring of support needs of affected children, may improve childhood development, but causality remains to be established.

**Key Points:**

- Previous studies suggest that prenatal infections, and the maternal immune activation that comes with them, are associated with child developmental outcomes. However, research to date has been based on infections data that is either self-reported or included infections diagnosed in hospital only.
- This study examined associations between prenatal infections, measured by both hospital-diagnosed infections and receipt of infection-related prescriptions, and child developmental concerns identified by health visitors at ages 6-8 weeks and 27-30 months.
- Hospital-diagnosed prenatal infections were consistently associated with developmental concerns. Maternal receipt of infection-related prescriptions during pregnancy were also associated with developmental concerns, but only those related to personal-social and emotional-behavioural-attention development.
- This suggests that prenatal infections, particularly severe infections, are associated with early childhood developmental outcomes.

## Introduction

Maternal health during pregnancy plays an important role in later childhood development. One important aspect of this is maternal infections, which occur commonly during pregnancy and can require treatment with prescription drugs, or hospital admission in more severe cases (Collier *et al*., 2009; WHO Global Maternal Sepsis Study Research Group, 2020). Animal models of prenatal maternal infection suggest that a mother’s immune response to an infectious agent (i.e. cytokine signalling, antibody production), which is known as maternal immune activation or MIA, creates a cascade of events that can impact upon fetal brain development (Garay *et al*., 2013; Massrali *et al*., 2022; Oskvig *et al*., 2012). Whilst more research is needed, human studies tend to be consistent with this (see Han *et al*., 2021).

Most studies examining the effects of prenatal infections on childhood development have focussed on links between prenatal maternal infections and formal diagnoses of neurodevelopmental conditions in children. For example, some research suggests prenatal infections are associated with diagnosis of autism in children, particularly among severe maternal infections involving hospitalisation as these involve higher levels of maternal immune activation (Atladottir *et al*., 2010; Jiang *et al*., 2016; Lee *et al*., 2015). Meanwhile, other research has examined potential links between prenatal infections and attention-deficit/hyperactivity disorder (ADHD), with mixed results (e.g. see Ginsberg *et al*., 2019; Mann and McDermott, 2011; Werenberg Dreier *et al*., 2016). Prenatal infections have also been linked to mental health conditions in adults such as schizophrenia and bipolar disorder (Cordeiro *et al*., 2015; Khandaker *et al*., 2013; Parboosing *et al*., 2013).

Whilst most studies focus on neurodevelopmental conditions or mental health conditions, there have also been a small number of studies examining the relationship between prenatal maternal infections and a broader range of early childhood developmental outcomes. One study, by Green *et al*. (2018), of linked administrative data from New South Wales, Australia, found hospital-diagnosed prenatal maternal infections to be associated with increased odds of social, emotional, physical, cognitive and communication developmental vulnerabilities in children at age 5. Similarly, a cohort study using data from Boston and Providence, USA, found prenatal bacterial infections to be associated with reduced cognitive performance at age 7 (Lee *et al*., 2020). However, other research, conducted by Hall *et al*. (2021) using data from a UK cohort study, found no association between hospital-recorded prenatal infections and childhood socioemotional developmental outcomes at age 3, but did find self-reported maternal infections to be associated with increased emotional problems. Finally, research by Kwok *et al*. (2022), which examined maternal infections in each trimester of pregnancy and associations with child cognitive outcomes, suggests that infections in the third trimester could have an effect on cognitive abilities later in childhood (although effect sizes were small).

There are, however, some important limitations to these existing pieces of research. Notably, they have tended to use data on prenatal infections that are either: (a) self-reported, and thus limited by issues relating to a lack of a strong verification that infections were present and by the retrospective nature of reporting (see Hall *et al*., 2021; Kwok *et al*., 2022), or (b) based on hospital records only, and thus limited by only including a minority of infections that are most severe (Green *et al*., 2018). The present study overcomes some of these limitations by making use of prenatal infections data from both hospital records and infection-related prescriptions. These were linked to data on birth records and routine child health reviews to form a large linked administrative health dataset covering children born in 2011-2015 in Greater Glasgow & Clyde, Scotland. This dataset was used to address the following aims:

**Aim 1:** To examine associations between prenatal maternal infections, measured as both hospital-diagnosed prenatal infections and receipt of infection-related prescription(s) during pregnancy, and having childhood developmental concerns identified by health visitors during early routine child health reviews.

**Aim 2:** To examine whether these associations vary by: (a) specific developmental outcome types (i.e. gross-motor-skills, hearing-communication, vision-social-awareness, personal-social, emotional-behavioural-attention, and speech-language-communication development), or (b) the trimester(s) in which infections occurred.

## Methods

### Data and participants

The linked administrative health dataset – made up of data from hospital records, birth records, prescriptions and child health reviews – included children born in the National Health Service (NHS) region of Greater Glasgow & Clyde between 2011 and 2015. This included all children who had linked maternal health records and complete records of child developmental outcomes from child health reviews conducted (routinely and universally) at ages 6-8 weeks and 27-30 months (for full details of child health reviews see Public Health Scotland, 2020; Scottish Government, 2015). In total, this gave a final sample of 55,856 children and their mothers. A flowchart providing full details of the sample selection procedure is provided in Figure 1.

**Figure 1.**
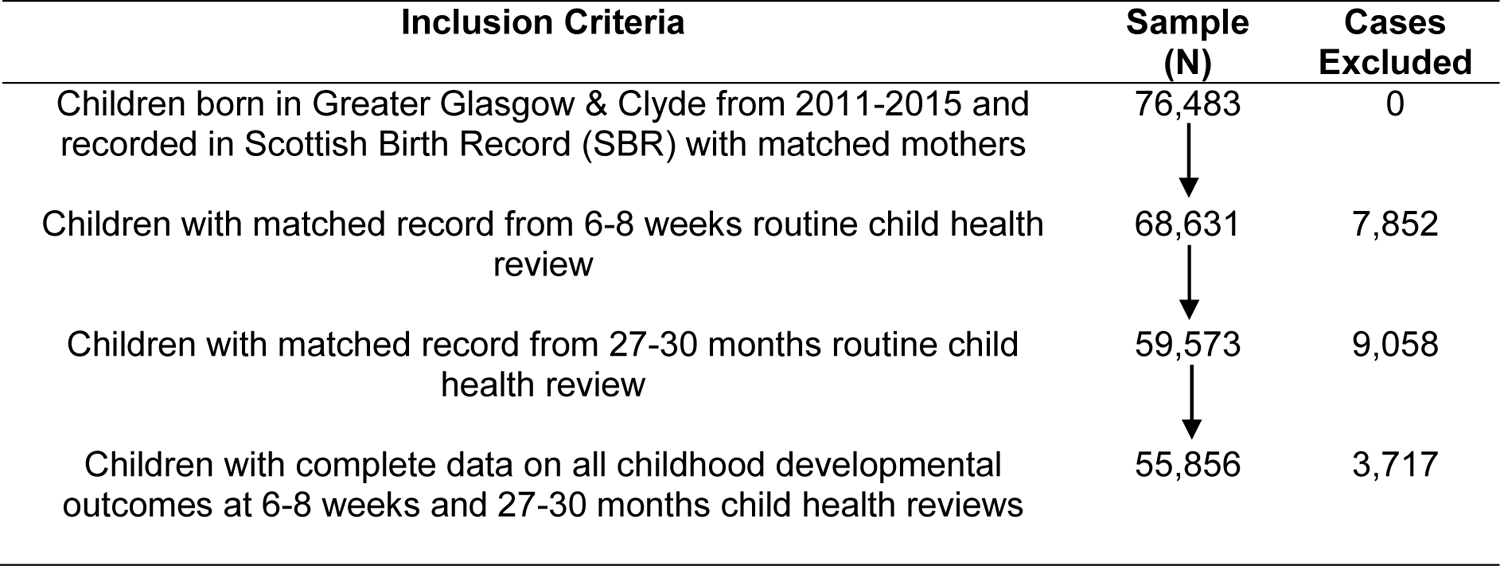
Flowchart of sample selection procedure

### Childhood developmental outcome measures

The first outcome variable was having any (i.e. at least one) childhood developmental concerns identified by health visitors during the 6-8 weeks or 27-30 months child health reviews of each child in the sample (coded: 0=‘No’, 1=‘Yes’). These could relate to either gross-motor-skills, hearing-communication or vision-social-awareness development measured at 6-8 weeks reviews, or personal-social, emotional-behavioural-attention, or speech-language-communication development measured at 27-30 months reviews. Developmental concerns were defined as cases where at least one of the above developmental observations were categorised by health visitors as ‘abnormal’, ‘a concern’ or ‘doubtful’ (i.e. suggestive of a possible abnormality, see: Playford et al., 2017) rather than as ‘normal’ or ‘no concerns’. Health visitors make these observations based on any/all of the following: (a) elicitation of parental concerns, (b) a structured observation of the child, and/or (c) the results of an ‘Ages and Stages’ questionnaire or other validated developmental assessment questionnaires (see: Scottish Government, 2015).

In addition, a further set of outcome variables was used in which the previous overall developmental concerns variable was disaggregated in order to indicate whether each child in the sample had developmental concerns identified that related specifically to: (a) gross-motor-skills, (b) hearing-communication, (c) vision-social-awareness, (d) personal-social, (e) emotional-behavioural-attention, and (f) speech-language-communication development. These outcomes were examined as six binary variables (each coded: 0=‘No’, 1=‘Yes’).

### Prenatal maternal infection measures

Two primary measures of prenatal maternal infection were used as explanatory variables in the analysis. The first was hospital-diagnosed prenatal infection(s), which indicates whether, for each mother in the sample, Scotland’s ‘general/acute inpatient and day case’ hospital records (see: Information Services Division Scotland, 2022d) or ‘maternity inpatient and day case’ hospital records (see: Information Services Division Scotland, 2022b) show the diagnosis of any maternal infection during the month of childbirth or in one of the nine preceding months (coded: 0=‘No’, 1=‘Yes’). Infections were identified using ICD10 codes. All ICD10 codes from hospital records were assessed and those indicating infection/inflammation were recorded as hospital-diagnosed prenatal infections. All postpartum/puerperal infections were excluded as although these were listed in maternity hospital records they would have occurred after delivery so are not prenatal infections. A full list of the ICD10 codes that were included is provided in Supporting Information Table S1.

The second primary measure used was receipt of infection-related prescription(s), which indicates whether, for each mother in the sample, Scotland’s Prescribing Information System (PIS, see: Information Services Division Scotland, 2022a) records show any receipt of prescriptions that are likely to have been for an infection during the month of their pregnancy or in one of the preceding nine months (coded: 0=‘No’, 1=‘Yes’). Like in existing literature making similar use of PIS data related to infections (e.g. Scott *et al*., 2018), infection-related prescriptions were defined as prescriptions included in British National Formulary (BNF) Chapter 5: Infections (British National Formulary, 2023). A full list of the drugs/prescriptions that were included is provided in Supporting Information Table S2. Notably, paracetamol prescriptions were not included in this measure, but were examined in later sensitivity analysis (outlined below).

In addition to the above primary explanatory variables, two further sets of explanatory variables were used in order to examine whether any potential associations between prenatal infections and childhood developmental outcomes varied depending on the trimester in which infections occurred. This is important to consider because fetal brain myelination does not begin until the second trimester of pregnancy (with further growth reinforcements occuring in trimester 3, see Cordeiro *et al*., 2015; Kwok *et al*., 2022). To measure this, the hospital-diagnosed prenatal infection(s) measure was disaggregated into: (a) hospital-diagnosed prenatal infection(s) in trimester 1, (b) hospital-diagnosed prenatal infection(s) in trimester 2, and (c) hospital-diagnosed prenatal infection(s) in trimester 3. Similarly, the receipt of infection-related prescriptions measure was disaggregated into: (a) receipt of infection-related prescription(s) during pregnancy trimester 1, (b) receipt of infection-related prescription(s) during pregnancy trimester 2, and (c) receipt of infection-related prescription(s) during pregnancy trimester 3.

Finally, one further additional explanatory variable, included as a sensitivity analysis, was receipt of any paracetamol prescription(s) during pregnancy. This indicates whether mothers of children in the sample received any paracetamol prescription during their pregnancy (coded: 0=‘No’, 1=‘Yes’). It was included an additional indicator of potential infection during pregnancy as clinical colleagues advised that paracetamol is frequently prescribed for infections during pregnancy.

### Confounders and covariates

The analysis also included a number of potential confounders and covariates as control variables. Maternal age and area-based deprivation were included as confounders, as these factors are known to be potentially associated with both maternal health during pregnancy and childhood health and developmental outcomes (Glick *et al*., 2021; Lean *et al*., 2017; Reijneveld *et al*., 2005; Vinikoor-Imler *et al*., 2011; Yasumitsu-Lovell *et al*., 2023). Data on maternal age comes from maternity hospital admissions data and was defined as maternal age at time of birth (coded: 0=≤19, 1=20-35, 2=≥36, i.e. in line with the ‘optimal’ (20-35) and ‘suboptimal’ (≤19/≥36) maternal age categorisations for neurodevelopment identified by previous research (Yasumitsu-Lovell *et al*., 2023)). Area-based deprivation comes from data on Scottish Index of Multiple Deprivation (SIMD), which was calculated from home postcodes and categorised into SIMD quintiles (coded: 1=most deprived, 2=more deprived, 3=medium deprived, 4=less deprived, and 5=least deprived).

In addition, sex of child, maternal history of mental health admissions and maternal prenatal smoking were included as covariates as they are potentially associated with childhood development (Green *et al*., 2018; National Advisory Council on Women and Girls, 2022; Wehby *et al*., 2011). Sex of child was defined as a binary variable (coded: 0=male, 1=female), with data on this coming from Scottish birth records. Data on maternal history of mental health admissions came from hospital records on mental health and inpatient and day case hospital admissions (see Information Services Division Scotland, 2022c). It was also defined as a binary variable (coded 0=no history of admissions, 1=history of admissions, i.e. one or more recorded admissions). Data was not available on less severe maternal mental health issues (i.e. those not requiring admission to hospital). Finally, maternal prenatal smoking was defined as a binary indicator of whether mothers smoked during pregnancy (coded: 0=no, 1=yes), and this information came from maternity hospital admissions data.

### Statistical analysis

The analysis used logistic regression models to measure associations between the prenatal maternal infection measures (i.e. hospital-diagnosed prenatal infections/receipt of infection-related prescriptions during pregnancy in the main analysis, and receipt of paracetamol prescriptions during pregnancy in the additional sensitivity analysis) and childhood developmental outcome measures (i.e. having any developmental concerns identified, and concerns identified specifically relating to gross-motor-skills, hearing-communication, vision-social-awareness, personal-social, emotional-behavioural-attention, and speech-language-communication development).

Modelling was conducted using a hierarchical approach, which involved (a) unadjusted models, (b) models which adjust for confounders only (i.e. maternal age and area-based deprivation), and (c) models which additionally adjust for covariates (i.e. sex of child, maternal history of mental health admissions and maternal prenatal smoking in addition to maternal age and area-based deprivation). The analysis was carried out as a complete case analysis as, whilst methods like multiple imputation can be used to deal with missing data on developmental outcomes from fully standardised measures like The Strength and Difficulties Questionnaire (SDQ), this is not appropriate when using outcomes related to health visitor judgements of developmental outcomes. All analysis was conducted using Stata/MP 16 software within Scotland’s National Safe Haven, with access to all datasets being facilitated by the electronic Data Research and Innovation Service (eDRIS) team at Public Health Scotland. The analysis plan for this study was preregistered using Open Science Framework (available here: https://osf.io/dx4vb), and the reporting in this study is consistent with ‘REporting of studies Conducted using Observational Routinely collected health Data’ (RECORD) guidelines (see Benchimol *et al*., 2015).

## Results

### Descriptive statistics

Descriptive statistics showing the characteristics of the sample, in terms of prevalence of childhood developmental outcomes and prenatal infections, as well as confounder/covariate characteristics, are provided in Table 1. Overall, 21.2% of children in the sample had at least one developmental concern identified by health visitors. Of these, concerns were more commonly identified for outcomes measured at 27-30 month health reviews, particularly those relating to speech-language-communication development (for which 13.1% of the sample had concerns identified) and emotional-behavioural-attention development (for which 10.5% of sample had concerns identified). Conversely, developmental concerns at 6-8 weeks child health reviews (i.e. gross-motor-skills, hearing-communication and vision-social-awareness development) were fairly rare, occurring for just 0.5%-1.8% of sample. Meanwhile, 5.1% of mothers in the sample had records of hospital-diagnosed infections, and 27.0% had records of receiving infection-related prescriptions (not including paracetamol), during pregnancy. With regards to confounders/covariates, it is noteworthy that maternal history of mental health admissions was fairly rare (1.6% of sample) and that the sample tended to be from more deprived SIMD quintiles, which reflects the fact that Glasgow has relatively high levels of deprivation (see Understanding Glasgow, 2020). In addition to Table 1, further descriptive statistics showing (a) variation in prenatal infection measures and confounders/covariates across childhood developmental outcomes, and (b) variation in childhood developmental outcomes and confounders/covariates across prenatal infection measures, are provided in Supporting Information Tables S3 and S4.

**Table 1.**
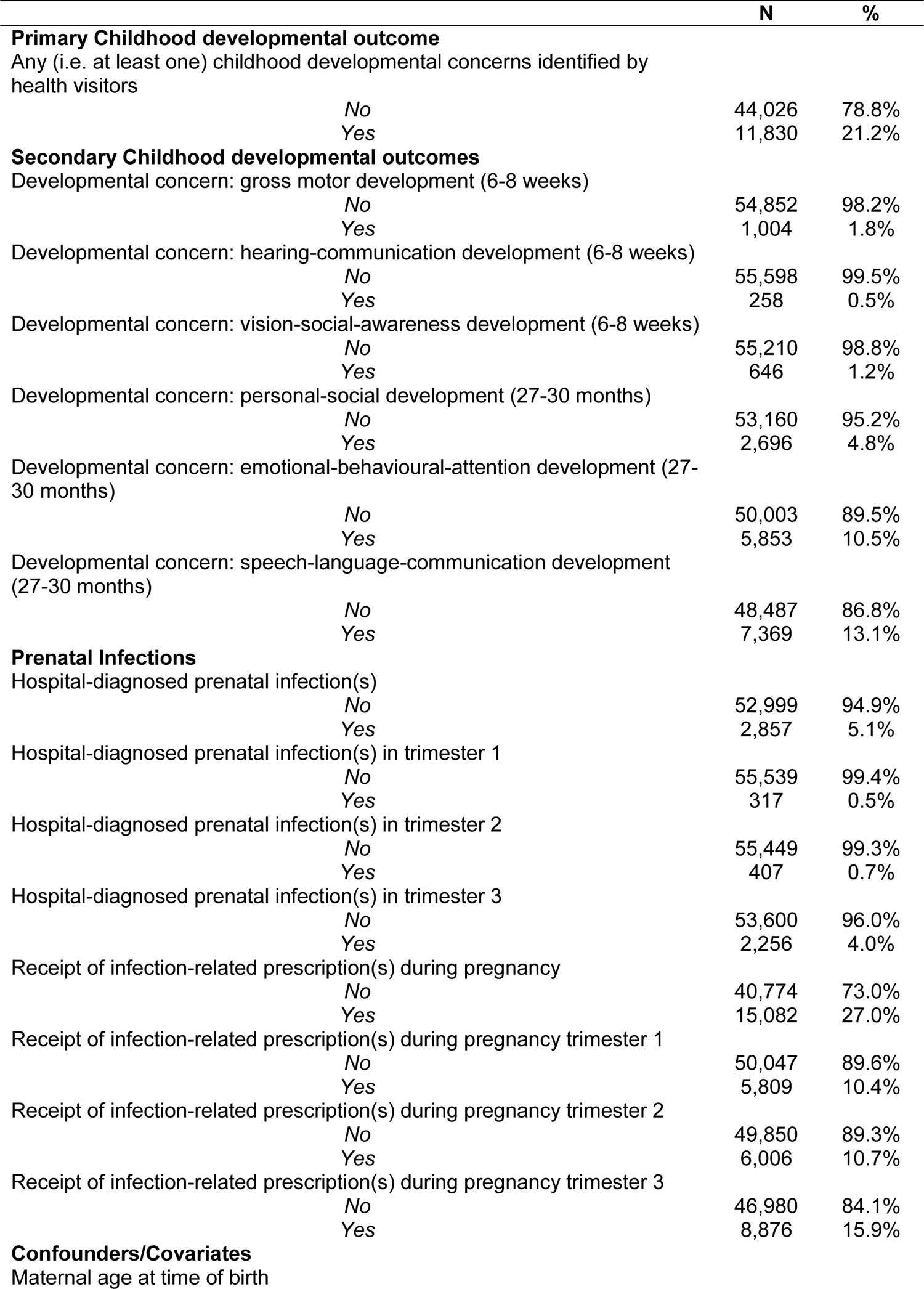
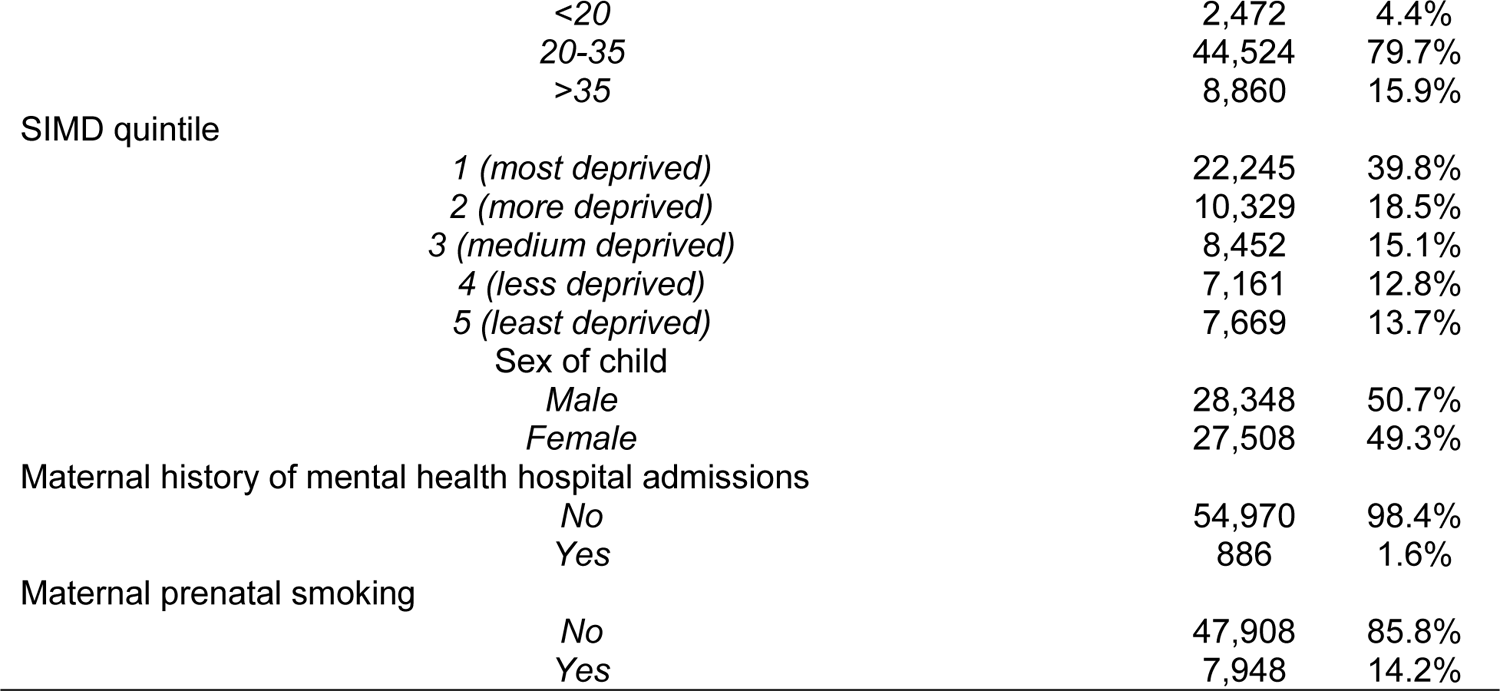
Descriptive statistics for childhood developmental outcomes, prenatal infections and confounders/covariates.

|  | N | % |
| --- | --- | --- |
| **Primary Childhood developmental outcome** |  |  |
| Any (i.e. at least one) childhood developmental concerns identified by health visitors |  |  |
| No | 44,026 | 78.8% |
| Yes | 11,830 | 21.2% |
| **Secondary Childhood developmental outcomes** |  |  |
| Developmental concern: gross motor development (6-8 weeks) |  |  |
| No | 54,852 | 98.2% |
| Yes | 1,004 | 1.8% |
| Developmental concern: hearing-communication development (6-8 weeks) |  |  |
| No | 55,598 | 99.5% |
| Yes | 258 | 0.5% |
| Developmental concern: vision-social-awareness development (6-8 weeks) |  |  |
| No | 55,210 | 98.8% |
| Yes | 646 | 1.2% |
| Developmental concern: personal-social development (27-30 months) |  |  |
| No | 53,160 | 95.2% |
| Yes | 2,696 | 4.8% |
| Developmental concern: emotional-behavioural-attention development (27-30 months) |  |  |
| No | 50,003 | 89.5% |
| Yes | 5,853 | 10.5% |
| Developmental concern: speech-language-communication development (27-30 months) |  |  |
| No | 48,487 | 86.8% |
| Yes | 7,369 | 13.1% |
| **Prenatal Infections** |  |  |
| Hospital-diagnosed prenatal infection(s) |  |  |
| No | 52,999 | 94.9% |
| Yes | 2,857 | 5.1% |
| Hospital-diagnosed prenatal infection(s) in trimester 1 |  |  |
| No | 55,539 | 99.4% |
| Yes | 317 | 0.5% |
| Hospital-diagnosed prenatal infection(s) in trimester 2 |  |  |
| No | 55,449 | 99.3% |
| Yes | 407 | 0.7% |
| Hospital-diagnosed prenatal infection(s) in trimester 3 |  |  |
| No | 53,600 | 96.0% |
| Yes | 2,256 | 4.0% |
| Receipt of infection-related prescription(s) during pregnancy |  |  |
| No | 40,774 | 73.0% |
| Yes | 15,082 | 27.0% |
| Receipt of infection-related prescription(s) during pregnancy trimester 1 |  |  |
| No | 50,047 | 89.6% |
| Yes | 5,809 | 10.4% |
| Receipt of infection-related prescription(s) during pregnancy trimester 2 |  |  |
| No | 49,850 | 89.3% |
| Yes | 6,006 | 10.7% |
| Receipt of infection-related prescription(s) during pregnancy trimester 3 |  |  |
| No | 46,980 | 84.1% |
| Yes | 8,876 | 15.9% |
| **Confounders/Covariates** |  |  |
| Maternal age at time of birth |  |  |

|  |  |  |  |
| --- | --- | --- | --- |
|  | <20 | 2,472 | 4.4% |
|  | 20-35 | 44,524 | 79.7% |
|  | >35 | 8,860 | 15.9% |
| SIMD quintile |  |  |  |
|  | 1 (*most deprived*) | 22,245 | 39.8% |
|  | 2 (*more deprived*) | 10,329 | 18.5% |
|  | 3 (*medium deprived*) | 8,452 | 15.1% |
|  | 4 (*less deprived*) | 7,161 | 12.8% |
|  | 5 (*least deprived*) | 7,669 | 13.7% |
|  | Sex of child |  |  |
|  | Male | 28,348 | 50.7% |
|  | Female | 27,508 | 49.3% |
| Maternal history of mental health hospital admissions |  |  |  |
|  | No | 54,970 | 98.4% |
|  | Yes | 886 | 1.6% |
| Maternal prenatal smoking |  |  |  |
|  | No | 47,908 | 85.8% |
|  | Yes | 7,948 | 14.2% |

### Prenatal infections and having at least one childhood developmental concern

Results showing associations between prenatal infections and having any (i.e. at least one) childhood developmental concerns identified by health visitors at 6-8 weeks or 27-30 months health reviews, whilst fully adjusting for confounders and covariates, are provided in Table 2. The results suggest that, after adjusting for confounders/covariates, hospital-diagnosed prenatal infections were associated with a 1.30 (95% CI: 1.19-1.42) increase in the odds of having at least one childhood developmental concern. However, there was no statistically significant relationship between receipt of infection-related prescriptions and having at least one childhood developmental concern after adjusting for confounders/covariates (OR: 1.03; 95% CI: 0.98-1.08).

**Table 2.** Odds ratios (95% CIs) for fully adjusted associations between prenatal infection(s) and having any (i.e. at least one) childhood developmental concern identified by health visitors.

|  | Having any (i.e. at least one)<br>childhood developmental concern<br>identified |
| --- | --- |
| **Hospital-diagnosed prenatal infection(s)** |  |
| [No] |  |
| Yes | 1.30*** (1.19-1.42) |
| **Receipt of infection-related prescription(s)**<br>**during pregnancy** |  |
| [No] |  |
| Yes | 1.03 (0.98-1.08) |

*Notes:* models fully adjust for maternal age at time of birth, SIMD quintile, sex of child, maternal history of mental health hospital admissions and maternal prenatal smoking. Reference categories are shown in square brackets. Childhood developmental concerns include those measured at both 6-8 weeks and 27-30 months routine child health visits. \*p<0.05, \*\*p<0.01, \*\*\*p<0.001.

Results of unadjusted and confounder adjusted models (as well as full details of the fully adjusted models, including odds ratios and 95% confidence intervals for confounders/covariates) are provided in Supporting Information Tables 5a and 5b. The results of models examining unadjusted and confounder adjusted associations between hospital-diagnosed prenatal infections and having at least one childhood developmental concern were similar to the results of the fully adjusted model, although associations were slightly stronger (OR: 1.43; 95% CI: 1.31-1.55 in unadjusted model and OR: 1.33; 95% CI: 1.22-1.45 in confounder adjusted model). Meanwhile, receipt of infection-related prescription(s) during pregnancy was associated with increased odds of having at least one childhood developmental concern in the unadjusted model (OR: 1.10; 95% CI: 1.05-1.16) but this was attenuated in the confounder adjusted model, which produced identical results to the fully adjusted model (OR: 1.03; 95% CI: 0.98-1.08).

### Prenatal infections and specific types of childhood developmental concerns

Results showing fully adjusted associations between prenatal infections and having specific types of childhood developmental concerns are shown in Table 3. They suggest that, after adjusting for confounders/covariates, hospital-diagnosed prenatal infections were associated with increased odds of childhood developmental concerns related to gross-motor-skills (OR: 1.30; 95% CI: 1.01-1.67) and vision-social-awareness (OR: 1.46; 95% CI: 1.08-1.96) development at age 6-8 weeks, and personal-social (OR: 1.34; 95% CI: 1.15-1.56), emotional-behaviour-attention (OR: 1.36; 95% CI: 1.22-1.52) and speech-language-communication (OR: 1.33; 95% CI: 1.20-1.47) development at age 27-30 months. However, no significant association was found between hospital-diagnosed prenatal infections and hearing-communication development at age 6-8 weeks.

**Table 3.**
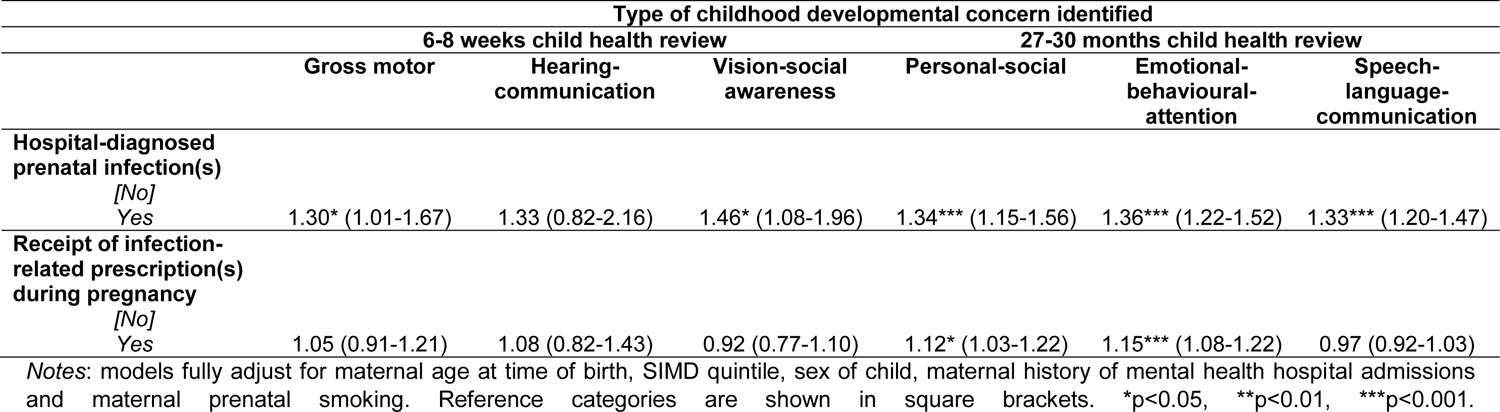
Odds ratios (95% CIs) for fully adjusted associations between prenatal infections and having specific types of childhood developmental concerns identified by health visitors.

With regards to infection-related prescriptions, the results suggest no significant association between receipt of infection-related prescriptions and most types of child developmental concerns examined. However, infection-related prescriptions were associated with a small increase in odds of concerns related specifically to personal-social (OR: 1.12; 95% CI: 1.03-1.22) and emotional-behaviour-attention (OR: 1.15; 95% CI: 1.08-1.22) development at age 27-30 months. In addition to the results of fully adjusted models, results of unadjusted and confounder adjusted models are provided in Supporting Information Tables 6a and 6b. These produced very similar results to those seen in fully adjusted models.

### Prenatal infections, by trimester, and having at least one childhood developmental concern

Results showing fully adjusted associations between prenatal infections, by trimester, and having at least one childhood developmental concern, are provided in Table 4. They highlight that the positive association between hospital-diagnosed prenatal infections and having at least one childhood developmental concern appeared to be specifically linked to infections occurring in trimesters 2 (OR: 1.34; 95% CI: 1.07-1.67) and 3 (OR: 1.33; 95% CI: 1.21-1.47). There was no significant association between hospital-diagnosed prenatal infections in trimester 1 and having at least one childhood developmental concern. In terms of infection-related prescriptions, there did not appear to be much variation in the relationship between their receipt and having childhood developmental concern(s) by trimester. Supporting Information Tables 7a and 7b provide full results of unadjusted and confounder adjusted associations between prenatal infections, by trimester, and having at least one childhood developmental concern. Overall, the results of the unadjusted and confounder adjusted models were similar to the fully adjusted models, but with slightly stronger associations.

**Table 4.** Odds ratios (95% CIs) for fully adjusted associations between prenatal infections, by trimester, and having any (i.e. at least one) childhood developmental concern identified by health visitors.

|  | Having any (i.e. at least one)<br>childhood developmental concern<br>identified |
| --- | --- |
| **Trimester 1 hospital-diagnosed prenatal infection(s)** |  |
| [No] |  |
| Yes | 1.11 (0.84-1.45) |
| **Trimester 2 hospital-diagnosed prenatal infection(s)** |  |
| [No] |  |
| Yes | 1.34* (1.07-1.67) |
| **Trimester 3 hospital-diagnosed prenatal infection(s)** |  |
| [No] |  |
| Yes | 1.33*** (1.21-1.47) |
| **Trimester 1 receipt of infection-related prescription(s) during pregnancy** |  |
| [No] |  |
| Yes | 1.09* (1.02-1.16) |
| **Trimester 2 receipt of infection-related prescription(s) during pregnancy** |  |
| [No] |  |
| Yes | 1.06* (1.00-1.14) |
| **Trimester 3 receipt of infection-related prescription(s) during pregnancy** |  |
| [No] |  |
| Yes | 1.04 (0.99-1.10) |

*Notes:* models fully adjust for maternal age at time of birth, SIMD quintile, sex of child, maternal history of mental health hospital admissions and maternal prenatal smoking. Reference categories are shown in square brackets. Childhood developmental concerns include those measured at both 6-8 weeks and 27-30 months routine child health visits. \*p<0.05, \*\*p<0.01, \*\*\*p<0.001.

### Sensitivity Analysis

The results of the sensitivity analysis, which modelled the relationship between receipt of paracetamol prescription(s) (used as an additional indicator of potential infections) and having at least one childhood developmental concern, are provided in Supporting Information Table S8. The results suggest that, after adjusting for confounders and covariates, receipt of paracetamol prescription(s) was associated with a small increase in the odds of having at least one childhood developmental concern (OR: 1.11; 95% CI: 1.04-1.19).

## Discussion

This study used linked administrative health data to examine the relationship between prenatal maternal infections, measured as both hospital-diagnosed infections and receipt of infection-related prescriptions, and early childhood developmental outcomes among children born in the NHS region of Greater Glasgow & Clyde, Scotland, between 2011 and 2015. The findings suggest that, after adjusting for measured confounders and covariates (i.e. maternal age at time of birth, SIMD quintile, sex of child, maternal history of mental health hospital admissions and maternal prenatal smoking), hospital-diagnosed prenatal infections were associated with increased odds of children having at least one developmental concern identified during 6-8 weeks or 27-30 months routine child health reviews. This relationship was consistent across almost all developmental outcome types measured, and appeared to be specifically linked to infections occurring in trimesters 2 and 3 of pregnancy. The findings also suggest that receipt of infection-related prescriptions during pregnancy (and indeed paracetamol prescriptions, which were included as an additional infection indicator) was associated with increased odds of children having at least one childhood developmental concern. However, this association between infection-related prescriptions and child development was attenuated after accounting for confounders and covariates. There were, however, increased odds of developmental concerns specifically related to personal-social and emotional-behavioural-attention development even after confounder and covariate adjustment. The relationship between receipt of infection-related prescriptions and developmental outcomes did not appear to vary by the trimester in which prescriptions were received.

Overall, these findings suggest that maternal immune activation, or some other mechanism arising from significant prenatal infections, is associated with increased likelihood of adverse childhood developmental outcomes. This is in line with what is observed in animal models of maternal prenatal infections (Garay *et al*., 2013; Massrali *et al*., 2022; Oskvig *et al*., 2012). In addition, the research findings of the present study suggest that prenatal infections diagnosed in hospital had a particularly clear association with developmental outcomes. This is in line with the results of some previous human studies such as Green *et al*. (2018), but not others e.g. Hall *et al*. (2021). This most likely reflects the fact that infections picked up in hospital will typically be more severe, and more severe infections will, in general, involve higher levels if maternal immune activation (see Hall *et al*., 2023; Jiang *et al*., 2016). In addition, the fact that this association appeared to be specifically related to infections in the second and third trimester of pregnancy most likely reflects the fact that fetal brain myelination occurs in the second and third trimester, and therefore this is the period in which maternal immune activation may have the greatest effect on prenatal brain development (see Cordeiro *et al*., 2015; Kwok *et al*., 2022).

One novel aspect of this study is that, whilst previous studies on prenatal infections and childhood development have relied on retrospectively self-reported survey data, or data on hospital-recorded infections only, the present study was able to also use data on receipt of infection-related prescriptions as an additional indicator of prenatal infection. The finding that receipt of infection-related prescriptions during pregnancy was associated with increased odds of developmental concerns related to personal-social and emotional-behavioural-attention development backs up previous research that found maternal self-reported prenatal infections to be associated with childhood emotional problems (Hall *et al*., 2021). However, the infection-related prescriptions measure used in the present study overcomes some of the limitations that this previous research had, in particular, limitations related to its self-reported data potentially lacking strong verification that an infection was present, and problems arising from the retrospective nature of its reporting (see Hall *et al*., 2021, p1647).

The key strength of this study was that it was able to make use of a large linked administrative health dataset, which included data on multiple indicators of prenatal infections, and a range of childhood developmental outcomes. Nevertheless, there are some limitations that must be noted. Firstly, whilst a range of confounders and covariates were controlled for, it is possible that there are additional confounding factors that were not observed in the dataset. For example, possible unobserved confounding mechanisms might be disruption of antenatal attachment caused by infection, or some genetic confounding linking susceptibility to infection, or to a general propensity to receive antibiotic prescriptions, and neurodevelopmental outcomes. In particular, given the subjective nature of antibiotic prescribing (see Borek *et al*., 2020; Public Health England, 2015), it is possible that unmeasured confounders with a genetic component (e.g. mild anxiety) might explain at least part of the observed association between infection-related prescriptions (specifically antibiotic prescriptions) during pregnancy and poorer child personal-social and emotional-behavioural-attention developmental outcomes (see Howie and Bigg, 1980). Also, given that childhood developmental outcomes are heritable, it is possible that parental behaviours related to their own neurodevelopmental outcomes could impact a mother’s likelihood of receiving a diagnosis of infection, e.g. by affecting their propensity to consult their GP about a possible infection.

In addition, there are also some limitations related to the administrative health dataset used in this study’s analysis. It is well documented that studies using administrative health data are often not fully representative of the target population due to missing data arising from people failing to interact with health services (e.g., due to barriers such as housing problems, living in remote areas or a wide range of other reasons), or due to inaccuracies in data linkage processes (see Harron *et al*., 2017; Shaw *et al*., 2022). In the case of the present study specifically, it was conducted as a complete case analysis, and participation in Scotland’s universal health visiting pathway (i.e. the routine child health reviews in which child developmental outcomes were measured) is known to be lower among those living in more deprived areas (Horne *et al*., 2021). This means that the study participants may not be completely representative of all children born in Greater Glasgow & Clyde during the analysis period. More broadly, Greater Glasgow & Clyde is also unlikely to be representative of Scotland (or the United Kingdom), as it is a mostly urban area and has relatively high levels of deprivation (Understanding Glasgow, 2020).

The analysis outlined in this study used hospital-diagnosed prenatal infections and receipt of infection-related prescriptions as the main prenatal infections indicators. As discussed above, this provides a fuller picture of infections than previous studies. However, each of these indicators comes with their own set of strengths and weaknesses. The strength of using hospital-diagnosed infections is that they provide a strong verification of the presence of an infection, as patients will have been thoroughly examined before a diagnosis is made. However, infections diagnosed in hospital likely includes only a small proportion of all infections, made up of the most severe cases only. Moreover, there can be nuances in the way hospital-diagnosed infections are coded and in how medical notes/records are interpreted by the medical reviewers entering ICD10 codes (Hashimoto *et al*., 2014). On the other hand, infection related prescriptions (and paracetamol prescriptions) cover a much larger number of prenatal infections, as PIS data covers all medicines that are prescribed and dispensed in the community (Information Services Division Scotland, 2022a). However, its accuracy in capturing infections may be affected by geographical or temporal variations in GP prescribing practices (see Devine *et al*., 2021; Guthrie *et al*., 2022; Pouwels *et al*., 2018b), and there have been known issues relating to inappropriate prescribing and overprescribing of antibiotics in the UK and elsewhere (e.g. see Pouwels *et al*., 2018a; Smieszek *et al*., 2018)..

To conclude, although this study has some limitations, by including data on both hospital-diagnosed infections and infection-related prescriptions it provides as full a picture as possible on prenatal infections. The study provides evidence that prenatal maternal infections, particularly those which are (a) hospital-diagnosed (and thus likely more severe), and (b) occurring in the second and third trimester of pregnancy, are associated with early childhood developmental outcomes in children. As this may be linked to the process of maternal immune activation affecting fetal brain development, prevention of prenatal infections in the second and third trimester of pregnancy, alongside monitoring of support needs of affected children, may improve childhood development. However, the findings from this study require confirmation in experimental study designs.

## Data Availability

The administrative health datasets used for this study are not publicly available. However, they can be accessed via successfully applying to the NHS Scotland Public Benefit and Privacy Panel for Health and Social Care

## Acknowledgements

Iain Hardie and Bonnie Auyeung were supported by the Economic and Social Research Council (ES/W001519/1) during the course of this work. Bonnie Auyeung was additionally supported by the European Union’s Horizon 2020 research and innovation programme under the Marie Skłodowska-Curie grant agreement No.813546, the Baily Thomas Charitable Fund TRUST/VC/AC/SG/469207686, the Data Driven Innovation. In addition, the authors would like to acknowledge the electronic Data Research and Innovation Service (eDRIS) team at Public Health Scotland for their support in obtaining approvals, the provisioning and linking of data and facilitating access to the National Safe Haven.

## Ethical Considerations

This project was approved by the research ethics committee for the School of Philosophy, Psychology and Language Sciences (PPLS) at the University of Edinburgh (reference: 190-1718/2). The project also received information governance approval from the NHS Scotland Public Benefit and Privacy Panel for Health and Social Care (HSC-PBPP, reference: 1617– 0314). Written consent from participants was not required or obtained. All data used in this study was housed within Scotland’s National Safe Haven, coordinated by the electronic Data Research and Innovation Service (eDRIS) team at Public Health Scotland. The data was only accessed by approved researchers.

