## Supporting Information for "Prenatal maternal infections and early childhood developmental outcomes: Analysis of linked administrative health data for Greater Glasgow & Clyde, Scotland"

**Table S1. List of ICD10 codes included in this study’s hospital-diagnosed prenatal infection**s definition

| **ICD10 Codes** | **Type of Infections Included** |
| --- | --- |
| A/B | virus infections; bacterial infections; sexually transmitted infections/diseases |
| G0/G531/G630/ G940 | meningitis; cranial nerve infections; polyneuropathy infections; hydrocephalus infections |
| H0/H1/H20/H22/H320/H440/H441/H451/H481 | eye infections |
| I30/I310/I311/I32/ I33/I38/I39/I40/  I41/I430/I520/I521/I681/I980/I981 | pericarditis; endocarditis; myocarditis; heart infections; artery infections; cardiovascular infections |
| J0/J1/J2?J31/J32/J35/J36/J37/  J39/J40/J41/J42/J440/J85/J86 | sinusitis; tonsillitis; laryngitis; common cold; influenza; pneumonia; respiratory infections; bronchitis; rhinitis; pharyngitis; peritonsillar abscess; pharynx infections; pulmonary disease with infection; gangrene/lung abscess; pyothorax |
| K04/K05/K102/K112/K113/K122/K140/K35/K61/K630/K65/K67/K75/K770/K81/K930 | periodontitis; pulpitis; gingivitis; necrosis; jaw inflammation/infection; salivary gland infection; cellulitis/mouth abscess; glossitis; appendicitis; anal infection/abscess; intestine abscess/infection; peritonitis; liver infection; cholecystitis; tuberculous of intestine |
| L0 | skin infections |
| M0 | sepctic arthitis |
| N080/N10/N11/N12/N22/N290/N291/N30/N330/N34/N390/N61/N7 | glomerular disorders involving infection; interstitial nephritis; urinary calculus infection; kidney infection; cystitis; urethritis; urinary tract infection unspecified; infection in breast; pelvic/vaginal/vulva infection or inflammation |
| O23/O98 | infections/infectious disease associated with pregnancy |
| R50 | fever |
| T880 | Infection following immunisation |

**Table S2. List of drugs/prescriptions included in this study’s** receipt of infection-related

| **Category of Drugs/**  **Prescriptions** | **Specific Drugs/Prescriptions Included** |
| --- | --- |
| Antibacterials (antibiotics) | -Penicillin (Amoxicillin, Ampicillin, Co-amoxiclav, Co-fluampicil, Flucloxacillin, Phenoxymethylpenicillin)  -Cephalosporins and other beta-lactams (Cefaclor, Cephalexin, Cefixime, Cefradine, Cefuroxime)  -Tetracyclines (Doxycycline, Lymecycline, Minocycline, Oxytetracycline, Clobetasone with Oxytetracycline and Nystatin, Tretracycline)  -Aminoglycosides (Gentamicin, Neomycin Sulfate, Betamethasone with Neomycin, Hydrocortisone with Neomycin, Triamcinolone Gramicidin Neoymycin and Nystatin, Dexamethasone with Neomycin and Polymyxin B)  -Macrolides (Azithromycin, Clarithromycin, Erythromycin, Erythromycin with Zinc Acetate, Isotretinoin with Erythromycin)  -Clindamycin and Lincomycin (clindamycin)  -Other Antibacterials (Chloramphenicol, Colistin, Fusidic Acid, Betamethason with Fusidic Acid, Hydrocortisone with Fusidic Acid, Vancomycin)  -Antituberculosis Drugs (Cycloserine, Ethambutol Hydrochloride, Rifampicin with Isoniazid, Rifampicin)  -Antileprotic Drugs (Dapsone)  -Metronidazole, Tinidazole and Ornidazole (Metronidazole, Tinidazole)  -Quinolones (Cifrofloxacin, Levofloxacin, Moxifloxacin, Norfloxacin, Ofloxacin)  -Urinary Tract Infection Drugs (Methenamine, Nitrofurantoin) |
| Antifungals | -Triazole Antifungals (Fluconazole, Fluconozole and Clotrimazole, Itraconazole)  -Imidazole Antifungals (Ketoconazole)  -Polyene Antifungals (Amphotericin, Nystatin)  -Other Antifungals (Griseofulvin) |
| Antivirals | -Herpesvirus Infections (Aciclovir, Famciclovir, Penciclovir, Valaciclovir)  -Viral Hepatitis (Enteclavir)  -Influenza (Amantadine Hydrochloride, Oseltamivir, Zanamivir) |
| Antiprotozoals | Antimalarial drugs (Mefloquine, Quinine, Proguanil Hydrochloride with Chloroquine Phosphate, Atovaquone with Proguanil Hydrochloride) |
| Antiprotozoals | -Drugs for Threadworms (Mebendazole, Piperazine with Senna) |

**Table S3. Descriptive statistics for prenatal infections and confounders/covariates by childhood developmental outcome(s)**

|  | **N (%)** | |
| --- | --- | --- |
|  | **Any (at least one) childhood developmental concerns** | |
|  | **No** | **Yes** |
| **Prenatal Infections** |  |  |
| Hospital-diagnosed prenatal infection(s) |  |  |
| *No* | 41,948 (79.2%) | 11,051 (20.8%) |
| *Yes* | 2,078 (72.7%) | 779 (27.3%) |
| Hospital-diagnosed prenatal infection(s) (trimester 1) |  |  |
| *No* | 43,788 (78.8%) | 11,751 (21.2%) |
| *Yes* | 238 (75.1%) | 79 (24.9%) |
| Hospital-diagnosed prenatal infection(s) (trimester 2) |  |  |
| *No* | 43,737 (78.9%) | 11,712 (21.1%) |
| *Yes* | 289 (71.0%) | 118 (29.0%) |
| Hospital-diagnosed prenatal infection(s) (trimester3) |  |  |
| *No* | 42,394 (79.1%) | 11,206 (20.9%) |
| *Yes* | 1,632 (72.3%) | 624 (27.7%) |
| Receipt of infection-related prescription(s) during pregnancy |  |  |
| *No* | 32,323 (79.3%) | 8,451 (20.7%) |
| *Yes* | 11,703 (77.6%) | 3,379 (22.4%) |
| Receipt of infection-related prescription(s) during pregnancy (trimester 1) |  |  |
| *No* | 39,606 (79.1%) | 10,441 (20.9%) |
| *Yes* | 4,420 (76.1%) | 1,389 (23.9%) |
| Receipt of infection-related prescription(s) during pregnancy (trimester 2) |  |  |
| *No* | 39,421 (79.1%) | 10,429 (20.9%) |
| *Yes* | 4,605 (76.7%) | 1,401 (23.3%) |
| Receipt of infection-related prescription(s) during pregnancy (trimester 3) |  |  |
| *No* | 37,159 (79.1%) | 9,821 (20.9%) |
| *Yes* | 6,867 (77.8%) | 2,009 (22.6%) |
| **Confounders/Covariates** |  |  |
| Maternal age at time of birth |  |  |
| *<20* | 1,680 (68.0%) | 792 (32.0%) |
| *20-35* | 35,095 (78.8%) | 9,429 (21.2%) |
| *>35* | 7,251 (81.8%) | 1,609 (18.2%) |
| SIMD quintile |  |  |
| *1 (most deprived)* | 16,444 (73.9%) | 5,801 (26.1%) |
| *2 (more deprived)* | 8,041 (77.9%) | 2,288 (22.1%) |
| *3 (medium deprived)* | 6,844 (81.0%) | 1,608 (19.0%) |
| *4 (less deprived)* | 6,018 (84.0%) | 1,143 (16.0%) |
| *5 (least deprived* | 6,679 (87.1%) | 990 (12.9%) |
| Sex of child |  |  |
| *Male* | 20,644 (72.8%) | 7,704 (27.2%) |
| *Female* | 23,382 (85.0%) | 4,126 (15.0%) |
| Maternal history of mental health hospital admissions |  |  |
| *No* | 43,425 (79.0%) | 11,545 (21.0%) |
| *Yes* | 601 (67.8%) | 285 (32.2%) |
| Maternal prenatal smoking |  |  |
| *No* | 38,607 (80.6%) | 9,301 (19.4%) |
| *Yes* | 5,419 (68.2%) | 2,529 (31.8%) |

**Table S4. Descriptive statistics for childhood developmental outcomes** and confounders/covariates by prenatal infections

|  | **N (%)** | | | |
| --- | --- | --- | --- | --- |
|  | **Hospital-recorded prenatal infection(s)** | | **Receipt of infection-related prescription(s) during pregnancy** | |
|  | **No** | **Yes** | **No** | **Yes** |
| **Primary Childhood developmental outcome** |  |  |  |  |
| Any (i.e. at least one) childhood developmental concerns identified by health visitors |  |  |  |  |
| *No* | 41,948 (95.4%) | 2,078 (4.7%) | 32,323 (73.4%) | 11,703 (26.6%) |
| *Yes* | 11,051 (93.4%) | 779 (6.6%) | 8,451 (71.4%) | 3,379 (28.6%) |
| **Secondary Childhood developmental outcomes** |  |  |  |  |
| Developmental concern: gross motor development (6-8 weeks) |  |  |  |  |
| *No* | 52,063 (94.9%) | 2,789 (5.1%) | 40,057 (73.0%) | 14,795 (27.0%) |
| *Yes* | 936 (93.2%) | 68 (6.8%) | 717 (71.4%) | 287 (28.6%) |
| Developmental concern: hearing-communication development (6-8 weeks) |  |  |  |  |
| *No* | 52,759 (94.9%) | 2,839 (5.1%) | 40,592 (73.0%) | 15,006 (27.0%) |
| *Yes* | 240 (93.0%) | 18 (6.9%) | 182 (70.5%) | 76 (29.5%) |
| Developmental concern: vision-social-awareness development (6-8 weeks) |  |  |  |  |
| *No* | 52,401 (94.9%) | 2,809 (5.1%) | 40,293 (73.0%) | 14,917 (27.0%) |
| *Yes* | 598 (92.6%) | 48 (7.4%) | 481 (74.5%) | 165 (25.6%) |
| Developmental concern: personal-social development (27-30 months) |  |  |  |  |
| *No* | 50,500 (95.0%) | 2,660 (5.0%) | 38,896 (73.2%) | 14,264 (26.8%) |
| *Yes* | 2,499 (92.7%) | 197 (7.3%) | 1,878 (69.7%) | 818 (30.3%) |
| Developmental concern: emotional-behavioural-attention development (27-30 months) |  |  |  |  |
| *No* | 47,573 (95.1%) | 2,430 (4.9%) | 36,748 (73.5%) | 13,255 (26.5%) |
| *Yes* | 5,426 (92.7%) | 427 (7.3%) | 4,026 (68.8%) | 1,827 (31.2%) |
| Developmental concern: speech-language-communication development (27-30 months) |  |  |  |  |
| *No* | 46,132 (95.1%) | 2,355 (4.9%) | 35,422 (73.1%) | 13,065 (26.9%) |
| *Yes* | 6,867 (93.2%) | 502 (6.8%) | 5,352 (72.6%) | 2,017 (27.4%) |
| **Confounders/Covariates** |  |  |  |  |
| Maternal age at time of birth |  |  |  |  |
| *<20* | 2,252 (91.1%) | 220 (8.9%) | 1,179 (47.7%) | 1,293 (52.3%) |
| *20-35* | 42,283 (95.0%) | 2,241 (5.0%) | 32,626 (73.3%) | 11,898 (26.7%) |
| *>35* | 8,464 (95.5%) | 396 (4.5%) | 6,969 (78.7%) | 1,891 (21.3%) |
| SIMD quintile |  |  |  |  |
| *1 (most deprived)* | 20,906 (94.0%) | 1,339 (6.0%) | 15,694 (70.6%) | 6,551 (29.4%) |
| *2 (more deprived)* | 9,778 (94.7%) | 551 (5.3%) | 7,423 (71.9%) | 2,906 (28.1%) |
| *3 (medium deprived)* | 8,052 (95.3%) | 400 (4.7%) | 6,197 (73.3%) | 2,255 (26.7%) |
| *4 (less deprived)* | 6,820 (95.2%) | 341 (4.8%) | 5,427 (75.8%) | 1,734 (24.2%) |
| *5 (least deprived* | 7,443 (97.1%) | 226 (2.9%) | 6,033 (78.7%) | 1,636 (21.3%) |
| Sex of child |  |  |  |  |
| *Male* | 26,878 (94.8%) | 1,470 (5.2%) | 20,753 (73.2%) | 7,595 (26.8%) |
| *Female* | 26,121 (95.0%) | 1,387 (5.0%) | 20,021 (72.8%) | 7,487 (27.2%) |
| Maternal history of mental health hospital admissions |  |  |  |  |
| *No* | 52,191 (94.9%) | 2,779 (5.1%) | 40,172 (73.1%) | 14,798 (26.9%) |
| *Yes* | 808 (91.2%) | 78 (8.8%) | 602 (67.9%) | 284 (32.1%) |
| Maternal prenatal smoking |  |  |  |  |
| *No* | 45,629 (95.2%) | 2,279 (4.8%) | 35,219 (73.5%) | 12,689 (26.5%) |
| *Yes* | 7,370 (92.7%) | 578 (7.3%) | 5,555 (73.0%) | 2,393 (30.1%) |

**Table S5a. Odds ratios (95% CIs) for unadjusted, confounder adjusted and fully adjusted associations between hospital-diagnosed prenatal infections and having any (i.e. at least one) childhood developmental concerns identified by health visitors**

|  | **Having any (i.e. at least one) childhood developmental concerns identified** | | |
| --- | --- | --- | --- |
|  | Unadjusted | Confounder adjusted | Fully adjusted |
| **Hospital-diagnosed prenatal infection(s)** |  |  |  |
| *[No]* |  |  |  |
| *Yes* | 1.43*** (1.31-1.55) | 1.33*** (1.22-1.45) | 1.30*** (1.19-1.42) |
| **Maternal age at time of birth** |  |  |  |
| *<20* |  | 1.55*** (1.42-1.69) | 1.49*** (1.35-1.63) |
| *[20-35]* |  |  |  |
| *>35* |  | 0.94* (0.88-1.00) | 0.94 (0.89-1.00) |
| **SIMD quintile** |  |  |  |
| *1 (most deprived)* |  | 1.46*** (1.28-1.56) | 1.39*** (1.30-1.48) |
| *2 (more deprived)* |  | 1.20*** (1.11-1.29) | 1.18*** (1.10-1.27) |
| *[3 (medium deprived)]* |  |  |  |
| *4 (less deprived)* |  | 0.81*** (0.75-0.89) | 0.84*** (0.77-0.91) |
| *5 (least deprived)* |  | 0.64*** (0.59-0.70) | 0.67*** (0.61-0.73) |
| **Sex of child** |  |  |  |
| *[Male]* |  |  |  |
| *Female* |  |  | 0.46*** (0.44-0.48) |
| **Maternal history of mental health hospital admissions** |  |  |  |
| *[No]* |  |  |  |
| *Yes* |  |  | 1.48*** (1.28-1.72) |
| **Maternal prenatal smoking** |  |  |  |
| *[No]* |  |  |  |
| *Yes* |  |  | 1.64*** (1.55-1.73) |

*Notes*: Reference categories are shown in square brackets. Childhood developmental outcomes include those measured at both 6-8 weeks and 27-30 months routine child health visits. *p<0.05, **p<0.01, ***p<0.001.

**Table S5b. Odds ratios (95% CIs) for unadjusted, confounder adjusted and fully adjusted associations between receipt of infection-related prescription(s) during pregnancy and having any (i.e. at least one) childhood developmental concerns identified by health visitors**

|  | **Having any (i.e. at least one) childhood developmental concerns identified** | | |
| --- | --- | --- | --- |
|  | Unadjusted | Confounder adjusted | Fully adjusted |
| **Receipt of infection-related prescription(s) during pregnancy** |  |  |  |
| *[No]* |  |  |  |
| *Yes* | 1.10*** (1.05-1.16) | 1.03 (0.98-1.08) | 1.03 (0.98-1.08) |
| **Maternal age at time of birth** |  |  |  |
| *<20* |  | 1.56*** (1.42-1.70) | 1.49*** (1.36-1.63) |
| *[20-35]* |  |  |  |
| *>35* |  | 0.94* (0.89-1.00 | 0.94 (0.89-1.00) |
| **SIMD quintile** |  |  |  |
| *1 (most deprived)* |  | 1.47*** (1.38-1.56) | 1.39*** (1.31-1.48) |
| *2 (more deprived)* |  | 1.20*** (1.12-1.29) | 1.18*** (1.10-1.27) |
| *[3 (medium deprived)]* |  |  |  |
| *4 (less deprived)* |  | 0.82*** (0.75-0.88) | 0.84*** (0.77-0.91) |
| *5 (least deprived)* |  | 0.64*** (0.59-0.70) | 0.67*** (0.61-0.73) |
| **Sex of child** |  |  |  |
| *[Male]* |  |  |  |
| *Female* |  |  | 0.46*** (0.44-0.48) |
| **Maternal history of mental health hospital admissions** |  |  |  |
| *[No]* |  |  |  |
| *Yes* |  |  | 1.49*** 1.29-1.73) |
| **Maternal prenatal smoking** |  |  |  |
| *[No]* |  |  |  |
| *Yes* |  |  | 1.64*** (1.55-1.74) |

*Notes*: Reference categories are shown in square brackets. Childhood developmental concerns include those measured at both 6-8 weeks and 27-30 months routine child health visits. *p<0.05, **p<0.01, ***p<0.001.

**Table S6a. Odds ratios (95% CIs) for unadjusted, confounder adjusted and fully adjusted associations between hospital-diagnosed prenatal infections and having specific types of childhood developmental concerns identified by health visitors**

|  | **Type of childhood developmental concerns identified** | | | | | | | | | | | | | | | | | |
| --- | --- | --- | --- | --- | --- | --- | --- | --- | --- | --- | --- | --- | --- | --- | --- | --- | --- | --- |
|  | **6-8 weeks child health review** | | | | | | | | | **27-30 months child health review** | | | | | | | | |
|  | **Gross motor** | | | **Hearing-communication** | | | **Vision-social awareness** | | | **Personal-social** | | | **Emotional-behavioural-attention** | | | **Speech-language-communication** | | |
|  | Unadjusted | Confounder adjusted | Fully adjusted | Unadjusted | Confounder adjusted | Fully adjusted | Unadjusted | Confounder adjusted | Fully adjusted | Unadjusted | Confounder adjusted | Fully adjusted | Unadjusted | Confounder adjusted | Fully adjusted | Unadjusted | Confounder adjusted | Fully adjusted |
| **Hospital-diagnosed prenatal infection(s)** |  |  |  |  |  |  |  |  |  |  |  |  |  |  |  |  |  |  |
| *[No]* |  |  |  |  |  |  |  |  |  |  |  |  |  |  |  |  |  |  |
| *Yes* | 1.35*  (1.06-1.74) | 1.30*  (1.01-1.67) | 1.30*  (1.01-1.67) | 1.39  (0.86-2.25) | 1.35  (0.83-2.20) | 1.33  (0.82-2.17) | 1.50**  (1.11-2.01) | 1.49**  (1.11-2.01) | 1.46*  (1.08-1.96) | 1.50***  (1.29-1.74) | 1.39***  (1.20-1.62) | 1.34***  (1.15-1.56) | 1.54***  (1.38-1.71) | 1.42***  (1.27-1.58) | 1.36***  (1.22-1.52) | 1.43***  (1.29-1.58) | 1.36***  (1.23-1.50) | 1.33***  (1.20-1.47) |
| **Maternal age at time of birth** |  |  |  |  |  |  |  |  |  |  |  |  |  |  |  |  |  |  |
| *<20* |  | 1.20  (0.91-1.58) | 1.22  (0.93-1.60) |  | 1.65*  (1.02-2.68) | 1.63*  (1.01-2.65) |  | 1.00  (0.68-1.46) | 0.99  (0.68-1.45) |  | 1.17  (0.99-1.38) | 1.10  (0.93-1.31) |  | 1.75***  (1.58-1.95) | 1.65***  (1.48-1.84) |  | 1.20**  (1.07-1.34) | 1.16*  (1.03-1.29) |
| *[20-35]* |  |  |  |  |  |  |  |  |  |  |  |  |  |  |  |  |  |  |
| *>35* |  | 1.18  (0.99-1.41) | 1.78  (0.99-1.40) |  | 1.01  (0.71-1.42) | 1.01  (0.71-1.42) |  | 1.15  (0.94-1.42) | 1.15  (0.93-1.42) |  | 0.93  (0.83-1.05) | 0.93  (0.83-1.05) |  | 0.83***  (0.76-0.90) | 0.83***  (0.76-0.91) |  | 0.94  (0.88-1.01) | 0.95  (0.88-1.02) |
| **SIMD quintile** |  |  |  |  |  |  |  |  |  |  |  |  |  |  |  |  |  |  |
| *1 (most deprived)* |  | 1.66*** (1.35-2.05) | 1.68***  (1.36-2.07) |  | 1.16  (0.79-1.71) | 1.14  (0.77-1.69) |  | 1.03  (0.81-1.30) | 0.99  (0.78-1.26) |  | 1.71***  (1.51-1.93) | 1,59***  (1.40-1.80) |  | 1.78***  (1.63-1.94) | 1.63***  (1.50-1.79) |  | 1.31***  (1.22-1.41) | 1.26***  (1.17-1.36) |
| *2 (more deprived)* |  | 1.55*** (1.23-1.96) | 1.56***  (1.24-1.97) |  | 1.36  (0.88-2.08) | 1.35  (0.88-2.07) |  | 1.05  (0.81-1.30) | 1.03  (0.79-1.35) |  | 1.30***  (1.13-1.50) | 1.28**  (1.11-1.47) |  | 1.32***  (1.20-1.46) | 1.29***  (1.17-1.43) |  | 1.11*  (1.02-1.21) | 1.09*  (1.01-1.20) |
| *[3 (medium deprived)]* |  |  |  |  |  |  |  |  |  |  |  |  |  |  |  |  |  |  |
| *4 (less deprived)* |  | 1.09  (0.83-1.43) | 1.09  (0.83-1.43) |  | 0.94  (0.57-1.57 | 0.95  (0.57-1.58) |  | 0.93  (0.69-1.26) | 0.95  (0.71-1.28) |  | 0.78**  (0.65-0.93) | 0.80*  (0.68-0.97) |  | 0.82**  (0.73-0.93) | 0.86*  (0.76-0.97) |  | 0.80***  (0.73-0.89) | 0.82***  (0.74-0.91) |
| *5 (least deprived)* |  | 0.99  (0.76-1.31) | 0.99  (0.75-1.30) |  | 1.09  (0.67-1.78) | 1.11  (0.68-1.80) |  | 0.91  (0.68-1.23) | 0.93  (0.69-1.26) |  | 0.57***  (0.47-0.68) | 0.60***  (0.49-0.72) |  | 0.65***  (0.57-0.73) | 0.69***  (0.61-0.79) |  | 0.61***  (0.55-0.68) | 0.63***  (0.56-0.70) |
| **Sex of child** |  |  |  |  |  |  |  |  |  |  |  |  |  |  |  |  |  |  |
| *[Male]* |  |  |  |  |  |  |  |  |  |  |  |  |  |  |  |  |  |  |
| *Female* |  |  | 0.76***  (0.67-0.87) |  |  | 0.80  (0.63-1.03) |  |  | 0.82*  (0.70-0.96) |  |  | 0.41***  (0.38-0.44) |  |  | 0.49***  (0.46-0.52) |  |  | 0.38***  (0.36-0.40) |
| **Maternal history of mental health hospital admissions** |  |  |  |  |  |  |  |  |  |  |  |  |  |  |  |  |  |  |
| *[No]* |  |  |  |  |  |  |  |  |  |  |  |  |  |  |  |  |  |  |
| *Yes* |  |  | 1.46  (0.96-2.21) |  |  | 1.41  (0.61-3.23) |  |  | 2.05**  (1.32-3.18) |  |  | 1.55***  (1.22-1.98) |  |  | 1.50***  (1.25-1.80) |  |  | 1.31**  (1.10-1.56) |
| **Maternal prenatal smoking** |  |  |  |  |  |  |  |  |  |  |  |  |  |  |  |  |  |  |
| *[No]* |  |  |  |  |  |  |  |  |  |  |  |  |  |  |  |  |  |  |
| *Yes* |  |  | 0.89  (0.74-1.07) |  |  | 1.15  (0.81-1.62) |  |  | 1.20  (0.96-1.48) |  |  | 1.72***  (1.56-1.89) |  |  | 1.94***  (1.81-2.08) |  |  | 1.45***  (1.35-1.56) |

*Notes*: Reference categories are shown in square brackets. *p<0.05, **p<0.01, ***p<0.001.

**Table S6b. Odds ratios (95% CIs) for unadjusted, confounder adjusted and fully adjusted associations between receipt of infection-related prescription(s) during pregnancy and having specific types of childhood developmental concerns identified by health visitors**

|  | **Type of childhood developmental concern identified** | | | | | | | | | | | | | | | | | |
| --- | --- | --- | --- | --- | --- | --- | --- | --- | --- | --- | --- | --- | --- | --- | --- | --- | --- | --- |
|  | **6-8 weeks child health review** | | | | | | | | | **27-30 months child health review** | | | | | | | | |
|  | **Gross motor** | | | **Hearing-communication** | | | **Vision-social awareness** | | | **Personal-social** | | | **Emotional-behavioural-attention** | | | **Speech-language-communication** | | |
|  | Unadjusted | Confounder adjusted | Fully adjusted | Unadjusted | Confounder adjusted | Fully adjusted | Unadjusted | Confounder adjusted | Fully adjusted | Unadjusted | Confounder adjusted | Fully adjusted | Unadjusted | Confounder adjusted | Fully adjusted | Unadjusted | Confounder adjusted | Fully adjusted |
| **Receipt of infection-related prescription(s) during pregnancy** |  |  |  |  |  |  |  |  |  |  |  |  |  |  |  |  |  |  |
| *[No]* |  |  |  |  |  |  |  |  |  |  |  |  |  |  |  |  |  |  |
| *Yes* | 1.08  (0.94-1.24) | 1.05  (0.91-1.21) | 1.05  (0.91-1.21 | 1.13  (0.86-1.48) | 1.08  (0.82-1.43) | 1.08  (0.82-1.43) | 0.92  (0.78-1.11) | 0.93  (0.77-1.11) | 0.92  (0.77-1.10) | 1.19***  (1.09-1.29) | 1.12**  (1.03-1.22) | 1.12*  (1.03-1.22) | 1.26***  (1.19-1.33) | 1.15***  (1.08-1.22) | 1.15***  (1.08-1.23) | 1.02  (0.97-1.08) | 0.97  (0.92-1.02) | 0.97  (0.92-1.03) |
| **Maternal age at time of birth** |  |  |  |  |  |  |  |  |  |  |  |  |  |  |  |  |  |  |
| *<20* |  | 1.20  (0.90-1.58) | 1.22  (0.92-1.60) |  | 1.64  (1.00-2.69) | 1.62  (0.99-2.65) |  | 1.04  (0.71-1.52) | 1.03  (0.70-1.51) |  | 1.15  (0.97-1.36) | 1.08  (0.91-1.28) |  | 1.72***  (1.54-1.91) | 1.61***  (1.44-1.80) |  | 1.22***  (1.09-1.36) | 1.18**  (1.05-1.32) |
| *[20-35]* |  |  |  |  |  |  |  |  |  |  |  |  |  |  |  |  |  |  |
| *>35* |  | 1.18  (0.99-1.41) | 1.18  (0.99-1.40) |  | 1.01  (0.71-1.43) | 1.01  (0.71-1.43) |  | 1.15  (0.93-1.42) | 1.14  (093-1.41) |  | 0.93  (0.83-1.05) | 0.94  (0.83-1.05) |  | 0.83***  (0.76-0.91) | 0.84***  (0.77-0.91) |  | 0.94  (0.88-1.01) | 0.95  (0.88-1.02) |
| **SIMD quintile** |  |  |  |  |  |  |  |  |  |  |  |  |  |  |  |  |  |  |
| *1 (most deprived)* |  | 1.67***  (1.35-2.05) | 1.68***  (1.36-2.08) |  | 1.17  (0.79-1.72) | 1.14  (0.78-1.69) |  | 1.03  (0.82-1.30 | 1.00  (0.79-1.36) |  | 1.71***  (1.51-1.93) | 1.59***  (1.40-1.80) |  | 1.78***  (1.63-1.94) | 1.64***  (1.50-1.79) |  | 1.32***  (1.22-1.42) | 1.26***  (1.17-1.36) |
| *2 (more deprived)* |  | 1.56***  (1.23-1.96) | 1.56***  (1.24-1.97) |  | 1.36  (0.89-2.08) | 1.35  (0.88-2/07) |  | 1.05  (0.80-1.37) | 1.04  (0.80-1.36) |  | 1.30***  (1.13-1.50) | 1.28**  (1.11-1.47) |  | 1.32***  (1.20-1.46) | 1.29***  (1.17-1.43) |  | 1.11*  (1.02-1.21) | 1.10*  (1.01-1.20) |
| *[3 (medium deprived)]* |  |  |  |  |  |  |  |  |  |  |  |  |  |  |  |  |  |  |
| *4 (less deprived)* |  | 1.09  (0.83-1.43) | 1.09  (0.83-1.43) |  | 0.95  (0.57-1.57) | 0.95  (0.57-1.59) |  | 0.93  (0.69-1.26) | 0.94  (0.70-1.27) |  | 0.78**  (0.66-0.93) | 0.81*  (0.68-0.96) |  | 0.83**  (0.73-0.93) | 0.86*  (0.76-0.97) |  | 0.80***  (0.73-0.89) | 0.82***  (0.74-0.91) |
| *5 (least deprived)* |  | 0.99  (0.75-1.30) | 0.99  (0.75-1.30) |  | 1.09  (0.66-1.77) | 1.11  (0.68-1.80) |  | 0.90  (0.67-1.22) | 0.92  (0.69-1.25) |  | 0.56***  (0.47-0.68) | 0.60***  (0.49-0.72) |  | 0.65***  (0.57-0.74) | 0.69***  (0.61-0.79) |  | 0.61***  (0.55-0.68) | 0.62***  (0.56-0.69) |
| **Sex of child** |  |  |  |  |  |  |  |  |  |  |  |  |  |  |  |  |  |  |
| *[Male]* |  |  |  |  |  |  |  |  |  |  |  |  |  |  |  |  |  |  |
| *Female* |  |  | 0.76***  (0.67-0.87) |  |  | 0.80  (0.62-3.26) |  |  | 0.82*  (0.70-0.96) |  |  | 0.41***  (0.38-0.45) |  |  | 0.49***  (0.46-0.52) |  |  | 0.38***  (0.36-0.40) |
| **Maternal history of mental health hospital admissions** |  |  |  |  |  |  |  |  |  |  |  |  |  |  |  |  |  |  |
| *[No]* |  |  |  |  |  |  |  |  |  |  |  |  |  |  |  |  |  |  |
| *Yes* |  |  | 1.47  (0.97-2.23) |  |  | 1.42  (0.62-3.26) |  |  | 2.09**  1.35-3.21) |  |  | 1.57***  (1.23-1.99) |  |  | 1.51***  (1.26-1.81) |  |  | 1.32**  (1.11-1.58) |
| **Maternal prenatal smoking** |  |  |  |  |  |  |  |  |  |  |  |  |  |  |  |  |  |  |
| *[No]* |  |  |  |  |  |  |  |  |  |  |  |  |  |  |  |  |  |  |
| *Yes* |  |  | 0.90  (0.75-1.07) |  |  | 1.15  (0.82-1.63) |  |  | 1.21  (0.97-1.50) |  |  | 1.73***  (1.57-1.90) |  |  | 1.95***  (1.82-2.09) |  |  | 1.45***  (1.36-1.55) |

*Notes*: Reference categories are shown in square brackets. *p<0.05, **p<0.01, ***p<0.001.

**Table S7a. Odds ratios (95% CIs) for unadjusted, confounder adjusted and fully adjusted associations between hospital-diagnosed prenatal infections, by trimester, and having any (i.e. at least one) childhood developmental concerns identified by health visitors**

|  | **Having any (i.e. at least one) childhood developmental concerns identified** | | | | | | | | |
| --- | --- | --- | --- | --- | --- | --- | --- | --- | --- |
|  | **Timing of hospital-diagnosed prenatal infection** | | | | | | | | |
|  | **Trimester 1** | | | **Trimester 2** | | | **Trimester 3** | | |
|  | Unadjusted | Confounder adjusted | Fully adjusted | Unadjusted | Confounder adjusted | Fully adjusted | Unadjusted | Confounder adjusted | Fully adjusted |
| **Hospital-diagnosed prenatal infection(s)** |  |  |  |  |  |  |  |  |  |
| *[No]* |  |  |  |  |  |  |  |  |  |
| *Yes* | 1.24  (0.95-1.60) | 1.13  (0.87-1.46) | 1.11  (0.84-1.45) | 1.53***  (1.23-1.89) | 1.39**  (1.12-1.72) | 1.34*  (1.07-1.67) | 1.45***  (1.32-1.59) | 1.37***  (1.24-1.50) | 1.33***  (1.21-1.47) |
| **Maternal age at time of birth** |  |  |  |  |  |  |  |  |  |
| *<20* |  | 1.56***  (1.43-1.71) |  |  | 1.56***  (1.43-1.70) | 1.50***  (1.37-1.64) |  | 1.55***  (1.42-1.69) | 1.49***  (1.36-1.64) |
| *[20-35]* |  |  |  |  |  |  |  |  |  |
| *>35* |  | 0.94*  (0.88-0.99) |  |  | 0.94*  (0.88-0.99) | 0.94  (0.89-1.00) |  | 0.94*  (0.88-0.99) | 0.94  (0.89-1.00) |
| **SIMD quintile** |  |  |  |  |  |  |  |  |  |
| *1 (most deprived)* |  | 1.47***  (1.38-1.56) | 1.39***  (1.31-1.48) |  | 1.47***  (1.38-1.56) | 1.39***  (1.31-1.48) |  | 1.47***  (1.38-1.56) | 1.39***  (1.31-1.48) |
| *2 (more deprived)* |  | 1.20***  (1.12-1.29) | 1.18***  (1.31-1.48) |  | 1.20***  (1.12-1.29) | 1.18***  (1.10-1.27) |  | 1.20***  (1.12-1.29) | 1.18***  (1.10-1.27) |
| *[3 (medium deprived)]* |  |  |  |  |  |  |  |  |  |
| *4 (less deprived)* |  | 0.81***  (0.75-0.89) | 0.84***  (0.76-0.91) |  | 0.82***  (0.75-0.89) | 0.83***  (0.77-0.90) |  | 0.82***  (0.75-0.89) | 0.83***  (0.76-0.91) |
| *5 (least deprived)* |  | 0.64***  (0.59-0.70) | 0.67***  (0.61-0.73) |  | 0.64***  (0.59-0.70) | 0.67***  (0.61-0.73) |  | 0.65***  (0.59-0.70) | 0.67***  (0.61-0.73) |
| **Sex of child** |  |  |  |  |  |  |  |  |  |
| *[Male]* |  |  |  |  |  |  |  |  |  |
| *Female* |  |  | 0.46***  (0.44-0.48) |  |  | 0.50***  (0.44-0.48) |  |  | 0.46***  (0.44-0.48) |
| **Maternal history of mental health hospital admissions** |  |  |  |  |  |  |  |  |  |
| *[No]* |  |  |  |  |  |  |  |  |  |
| *Yes* |  |  | 1.50***  (1.29-1.73) |  |  | 1.50***  (1.29-1.73) |  |  | 1.48***  (1.28-1.73) |
| **Maternal prenatal smoking** |  |  |  |  |  |  |  |  |  |
| *[No]* |  |  |  |  |  |  |  |  |  |
| *Yes* |  |  | 1.64***  (1.56-1.74) |  |  | 1.64***  (1.55-1.74) |  |  | 1.64***  (1.55-1.73) |

*Notes*: Reference categories are shown in square brackets. Childhood developmental concerns include those measured at both 6-8 weeks and 27-30 months routine child health visits. *p<0.05, **p<0.01, ***p<0.001.

**Table S7b. Odds ratios (95% CIs) for unadjusted, confounder adjusted and fully adjusted associations between receipt of infection-related prescription(s), by trimester, and having any (i.e. at least one) childhood developmental concerns identified by health visitors**

|  | **Having any (i.e. at least one) childhood developmental concerns identified** | | | | | | | | |
| --- | --- | --- | --- | --- | --- | --- | --- | --- | --- |
|  | **Timing of hospital-diagnosed prenatal infection** | | | | | | | | |
|  | **Trimester 1** | | | **Trimester 2** | | | **Trimester 3** | | |
|  | Unadjusted | Confounder adjusted | Fully adjusted | Unadjusted | Confounder adjusted | Fully adjusted | Unadjusted | Confounder adjusted | Fully adjusted |
| **Receipt of infection-related prescription(s)** |  |  |  |  |  |  |  |  |  |
| *[No]* |  |  |  |  |  |  |  |  |  |
| *Yes* | 1.20***  (1.12-1.27) | 1.10**  (1.03-1.18) | 1.09*  (1.02-1.16) | 1.15***  (1.08-1.23) | 1.06  (0.99-1.14) | 1.06*  (1.00-1.14) | 1.11***  (1.05-1.17) | 1.05  (0.99-1.11) | 1.04  (0.99-1.10) |
| **Maternal age at time of birth** |  |  |  |  |  |  |  |  |  |
| *<20* |  | 1.54***  (1.41-1.69) | 1.49***  (1.35-1.63) |  | 1.55***  (1.42-1.70) | 1.49***  (1.36-1.63) |  | 1.55***  (1.42-1.70) | 1.49***  (1.36-1.64) |
| *[20-35]* |  |  |  |  |  |  |  |  |  |
| *>35* |  | 0.94*  (0.89-0.99) | 0.94  (0.89-1.00) |  | 0.94*  (0.89-0.99) | 0.94  (0.89-1.00) |  | 0.94*  (0.89-0.99) | 0.94*  (0.89-0.99) |
| **SIMD quintile** |  |  |  |  |  |  |  |  |  |
| *1 (most deprived)* |  | 1.47***  (1.38-1.56) | 1.39***  (1.30-1.48) |  | 1.47***  (1.38-1.56) | 1.39***  (1.31-1.48) |  | 1.47***  (1.38-1.56) | 1.39***  (1.31-1.48) |
| *2 (more deprived)* |  | 1.20***  (1.12-1.29) | 1.18***  (1.10-1.27) |  | 1.20***  (1.12-1.29) | 1.18***  (1.10-1.27) |  | 1.20***  (1.12-1.29) | 1.18***  (1.10-1.27) |
| *[3 (medium deprived)]* |  |  |  |  |  |  |  |  |  |
| *4 (less deprived)* |  | 0.82***  (0.75-0.88) | 0.84***  (0.77-0.90) |  | 0.82***  (0.75-0.88) | 0.84***  (0.77-0.90) |  | 0.82***  (0.75-0.89) | 0.84***  (0.77-0.91) |
| *5 (least deprived)* |  | 0.64***  (0.59-0.70) | 0.67***  (0.61-0.73) |  | 0.64***  (0.59-0.70) | 0.67***  (0.61-0.72) |  | 0.64***  (0.59-0.70) | 0.67***  (0.61-0.73) |
| **Sex of child** |  |  |  |  |  |  |  |  |  |
| *[Male]* |  |  |  |  |  |  |  |  |  |
| *Female* |  |  | 0.46***  (0.44-0.48) |  |  | 0.46***  (0.44-0.48) |  |  | 0.46***  (0.44-0.48) |
| **Maternal history of mental health hospital admissions** |  |  |  |  |  |  |  |  |  |
| *[No]* |  |  |  |  |  |  |  |  |  |
| *Yes* |  |  | 1.49***  (1.28-1.73) |  |  | 1.49***  (1.29-1.73) |  |  | 1.50***  (1.29-1.73) |
| **Maternal prenatal smoking** |  |  |  |  |  |  |  |  |  |
| *[No]* |  |  |  |  |  |  |  |  |  |
| *Yes* |  |  | 1.64***  (1.55-1.74) |  |  | 1.64***  (1.56-1.74) |  |  | 1.64***  (1.55-1.74) |

*Notes*: Reference categories are shown in square brackets. *p<0.05, **p<0.01, ***p<0.001.

**Table S8. Odds ratios (95% CIs) for unadjusted, confounder adjusted and fully adjusted associations between receipt of paracetamol prescription(s) during pregnancy and having any (i.e. at least one) childhood developmental concerns identified by health viusitors**

|  | **Having any (i.e. at least one) childhood developmental concerns identified** | | |
| --- | --- | --- | --- |
|  | Unadjusted | Confounder adjusted | Fully adjusted |
| **Receipt of paracetamol prescription(s) during pregnancy** |  |  |  |
| *[No]* |  |  |  |
| *Yes* | 1.24***  (1.16-1.33) | 1.12**  (1.05-1.20) | 1.11**  (1.04-1.19) |
| **Maternal age at time of birth** |  |  |  |
| *<20* |  | 1.55***  (1.41-1.69) | 1.49***  (1.36-1.63) |
| *[20-35]* |  |  |  |
| *>35* |  | 0.94*  (0.89-0.99) | 0.94  (0.89-1.00) |
| **SIMD quintile** |  |  |  |
| *1 (most deprived)* |  | 1.46***  (1.37-1.56) | 1.39***  (1.30-1.48) |
| *2 (more deprived)* |  | 1.20***  (1.11-1.29) | 1.18***  (1.10-1.27) |
| *[3 (medium deprived)]* |  |  |  |
| *4 (less deprived)* |  | 0.82***  (0.75-0.89) | 0.84***  (0.77-0.91) |
| *5 (least deprived)* |  | 0.64***  (0.59-0.70) | 0.67***  (0.61-0.72) |
| **Sex of child** |  |  |  |
| *[Male]* |  |  | 0.46***  (0.44-0.48) |
| *Female* |  |  |  |
| **Maternal history of mental health hospital admissions** |  |  |  |
| *[No]* |  |  |  |
| *Yes* |  |  | 1.49***  1.29-1.72) |
| **Maternal prenatal smoking** |  |  |  |
| *[No]* |  |  |  |
| *Yes* |  |  | 1.64***  (1.55-1.74) |

Notes: Reference categories are shown in square brackets. *p<0.05, **p<0.01, ***p<0.001.
